# Antimicrobial Stewardship in Pluralistic Health Systems: Why Local Evidence Must Guide Implementation

**DOI:** 10.64898/2026.09.24.26363886

**Authors:** Dipasri Konar, Girish Patil, Itismita Pradhan, Devesh Singh, Raman Kataria

**Affiliations:** Jan Swasthya Sahyog, Ganiyari, Bilaspur, Chhattisgarh, India

**Keywords:** antimicrobial resistance, antimicrobial stewardship, behavioral stewardship, WHO/INRUD, rational drug use, informal providers, injection use, primary care, India

## Abstract

**Background:** Antimicrobial resistance (AMR) strategies frequently emphasize provider knowledge and public awareness, but prescribing behavior can also reflect practice routines, information sources, and facility context. We examined these relationships in a pluralistic primary-care system in three districts of Chhattisgarh, India.

**Methods:** We analysed three independently sampled cross-sectional components: a provider knowledge, attitudes, and practices (KAP) survey (n=171), a WHO/International Network for Rational Use of Drugs (WHO/INRUD) prescribing assessment (600 encounters from 20 government facilities), and 371 patient exit interviews. For providers, proportional-odds logistic regression modelled ordered cold/sore-throat antibiotic provision (none, some, most, all), adjusting for provider cadre, work setting, and years in practice; binary any-versus-none models were sensitivity analyses. We summarized WHO/INRUD indicators overall and by facility, and examined exploratory encounter-level antibiotic correlates with unadjusted and facility-adjusted models. We analysed patient satisfaction using multivariable binary logistic regression and perceived clarity using proportional-odds logistic regression.

**Results:** Provider AMR awareness was 91.4% (149/163), 95.8% (160/167) agreed that antibiotics were overused nationally, and 58.3% (95/163) reported guideline familiarity. Guideline familiarity was not associated with lower cold/sore-throat antibiotic frequency (adjusted common OR 1.13, 95% CI 0.57-2.24; p=0.725). Each additional 5 years in practice was associated with greater odds of being in a higher prescribing-frequency category (adjusted common OR 1.36, 95% CI 1.09-1.70; p=0.006 before multiplicity correction). Medical-representative literature was a routine information source for 39.6% (65/164); its association with prescribing frequency was directionally positive but imprecise (adjusted common OR 1.89, 95% CI 0.88-4.07; p=0.104). In government facilities, 54.8% (329/600) encounters included an antibiotic, ranging from 16.7% to 80.0% by facility. Antibiotic-containing encounters had more drugs (2.88 vs 2.47; p<0.001), lower generic proportions (73.6% vs 80.7%; p=0.003), lower Essential Drugs List concordance (48.9% vs 63.8%; p<0.001), and more co-prescribed injections (15.2% vs 7.7%; p=0.007); several associations attenuated after accounting for facility. In exit interviews, 92.3% (131/142) of informal-provider encounters involved a reported injection, and informal providers accounted for 72.4% (131/181) of all reported injections. Clear/very clear instructions were strongly associated with satisfaction (adjusted OR 13.66, 95% CI 6.73-27.71; p<0.001; n=369).

**Conclusions:** In these three districts, high provider AMR awareness coexisted with prescribing patterns not explained by awareness or guideline familiarity alone. The findings support prospective evaluation of point-of-care decision support, independent drug information, facility audit and feedback, targeted informal-provider engagement, and communication-focused stewardship. Generalization beyond the study districts requires caution.

## 1. INTRODUCTION

Antimicrobial resistance (AMR) threatens global health security, universal health coverage, and sustainable development. International policy frameworks, including the World Health Organization (WHO) Global Action Plan on AMR, emphasize education, training, surveillance, and public awareness. Evidence from India and other health systems also indicates that knowledge dissemination alone may be insufficient to change antimicrobial prescribing when clinical uncertainty, social norms, workflow, and incentives remain unaddressed. [1–5]

In India, primary care is delivered through a pluralistic system in which modern-medicine practitioners work alongside practitioners of AYUSH systems (Ayurveda, Yoga and Naturopathy, Unani, Siddha and Homoeopathy), allied health workers, and informal providers. Treatment decisions in such mixed delivery systems may be influenced by diagnostic uncertainty, pharmaceutical marketing, local practice norms, payment arrangements, and perceived patient expectations. [6–10]

To address these drivers, antimicrobial stewardship (AMS) must evolve from passive guideline dissemination toward behavioral stewardship—interventions that directly target clinical choice architecture, facility norms, information environments, and clinician-patient communication. [2–5]

Using Bilaspur, Raigarh, and Sarguja districts of Chhattisgarh, India, as the study setting, we examined whether provider knowledge, facility prescribing patterns, and patient-reported treatment experience identify potentially modifiable stewardship targets. The purpose was not to infer that these districts represent Chhattisgarh, India, or other countries, but to evaluate a system-level triangulation approach within the populations actually sampled.

## 2. METHODS

### 2.1 Study Design and Setting

This multi-component cross-sectional study was conducted in Bilaspur, Raigarh, and Sarguja districts of Chhattisgarh, India, encompassing rural and urban primary care settings. The three sampling streams were independently recruited: Provider KAP data were collected from April 11 to September 30, 2022; patient exit-interview data were collected from September 17, 2022, to February 9, 2023; and the WHO/INRUD audit was conducted from September 12, 2022, to February 9, 2023. The components were not conducted fully concurrently.

1. **Provider KAP Component:** Evaluated provider awareness, guideline knowledge, information reliance, and self-reported practice.
2. **WHO/INRUD Prescribing Audit:** Conducted in government primary healthcare facilities to measure objective drug-use metrics.
3. **Patient Exit-Interview Component:** Sampled patient/caregiver experiences across government, NGO, private, and informal healthcare settings.

The patient exit-interview component was sampled independently from the provider KAP component and was not restricted to the same facilities or providers. This design was intended to reduce facility-specific selection and common-setting effects and to provide an independent assessment of patient treatment experience across the broader local healthcare market. The consequence is that provider KAP responses cannot be linked directly to the treatment experiences reported in exit interviews; cross-component comparisons therefore represent system-level triangulation rather than provider-level associations.

**Table 1.**
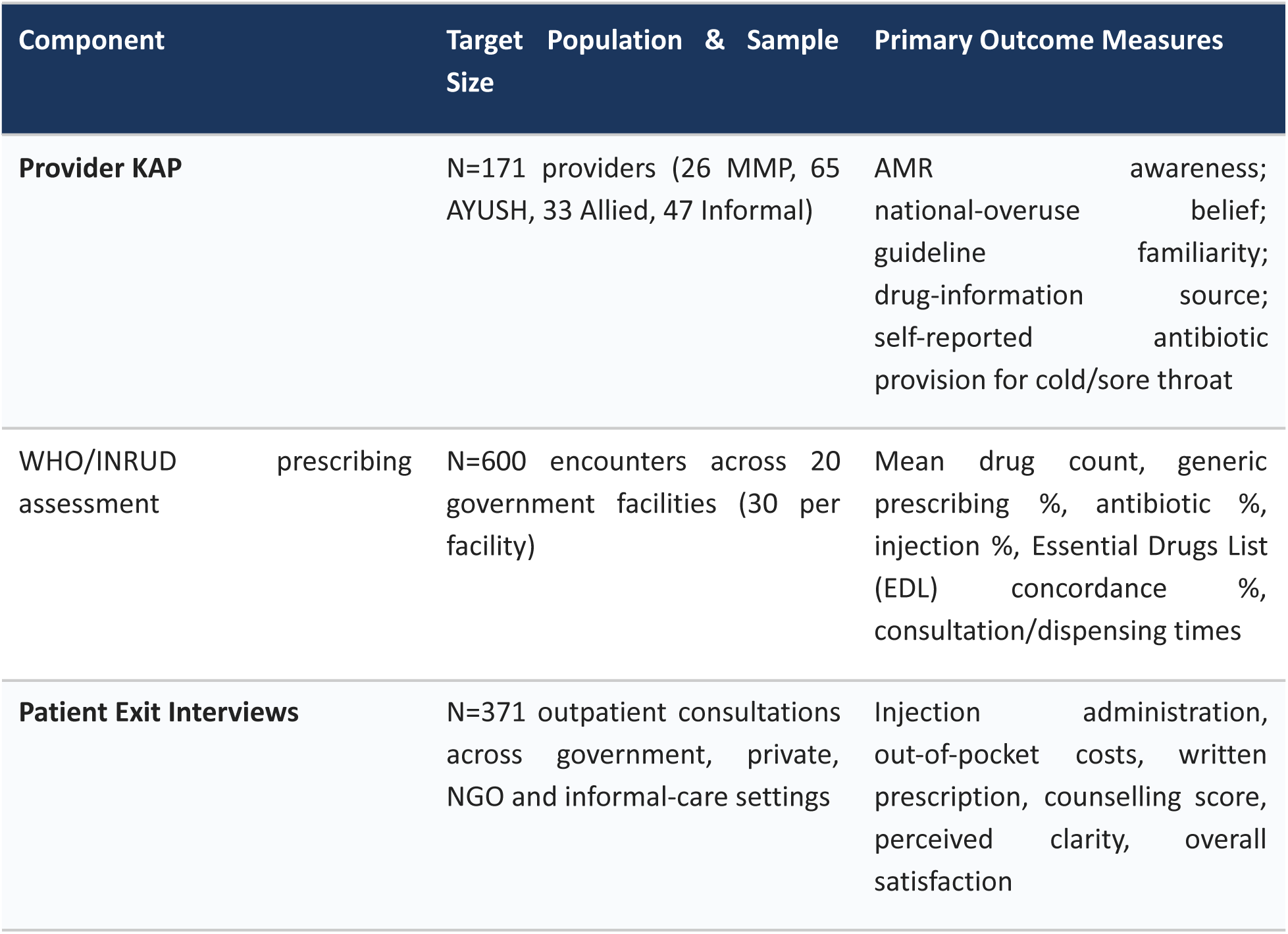
Overview of Study Components and Analytic Samples.

### 2.2 Provider KAP Survey

Eligible providers had at least 6 months of clinical practice in the study districts. Provider cadre was classified as: modern-medicine practitioners (MMPs), including Bachelor of Medicine, Bachelor of Surgery (MBBS), Doctor of Medicine (MD), Master of Surgery (MS), or postgraduate diploma qualifications; AYUSH practitioners, including Bachelor of Ayurvedic Medicine and Surgery (BAMS) and Bachelor of Homoeopathic Medicine and Surgery (BHMS); allied health professionals, including Auxiliary Nurse Midwives (ANMs), General Nursing and Midwifery (GNM) personnel, dentists, village/community health workers, and Mitanins; and informal practitioners who did not meet those definitions. The structured questionnaire was administered in Hindi and English and assessed AMR recognition, belief in national antibiotic overuse, guideline familiarity, drug-information sources, and self-reported antibiotic provision for cold or sore throat.

### 2.3 WHO/INRUD Prescribing Audit

The WHO/International Network for Rational Use of Drugs (WHO/INRUD) prescribing assessment used the standard core drug-use indicators as defined in WHO/DAP/93.1. [11] At each of 20 government primary health-care facilities, 30 outpatient encounters were assessed (N=600). The encounter was the unit of analysis. Prescribing indicators included mean number of medicines per encounter, percentage prescribed by generic name, percentage of encounters containing at least one antibiotic, percentage containing at least one injection, and percentage of medicines from the Essential Drugs List (EDL). Patient-care indicators, also defined according to WHO/DAP/93.1, included average consultation time, average dispensing time, percentage of prescribed medicines actually dispensed, percentage of dispensed medicines adequately labelled, and patient knowledge of the correct dosage. [11]

Facility sector describes where care was delivered and was coded as government, private, or non-governmental organization (NGO). Provider cadre describes the type of practitioner and was coded as MMP, AYUSH, allied, or informal. These variables were kept distinct. The WHO/INRUD prescribing assessment included government facilities only. In exit-interview analyses, facility sector was used for expenditure and satisfaction comparisons, whereas provider type was used for injection and communication comparisons.

### 2.4 Patient Exit Interviews

Consecutive patients or caregivers were interviewed immediately following outpatient visits across public, NGO, private, and informal settings, irrespective of whether medications were prescribed. Data included out-of-pocket expenditure, written prescription receipt, injection exposure, a 5-point counselling score, perceived clarity of instructions, and overall satisfaction. Facility sector and the cadre/type of the consulted provider were recorded as separate classifications.

### 2.5 Provider KAP Measures and Quality Control

Provider outcomes were defined from prespecified questionnaire items. Descriptive outcomes were: AMR awareness; agreement that antibiotics are overused in India; familiarity with antibiotic-use guidelines; routine use of medical-representative literature as a drug-information source; and frequency of antibiotic provision for cold or sore throat. Cold/sore-throat practice was ordered as 0=none, 1=some patients, 2=most patients, and 3=all patients. The hypothesis-directed exposures were guideline familiarity (yes/no) and routine use of medical-representative literature (yes/no). Covariates were provider cadre, work setting, and years in practice; gender was additionally included in the broader separated-domain models.

Cold/sore-throat presentations were selected as a pragmatic tracer of antibiotic-prescribing behavior because most uncomplicated upper-respiratory presentations are viral or self-limiting and are a well-recognized context for unnecessary antibiotic use. WHO AWaRe guidance emphasizes that the majority of upper respiratory tract infections generally should not be treated with antibiotics, while recognizing that antibiotics may be indicated in selected bacterial or severe presentations. Accordingly, the questionnaire item was used as a behavioral indicator of prescribing tendency and not as a diagnosis-specific measure of inappropriate prescribing. [12]

The English and Hindi provider questionnaire underwent forward and back translation, followed by review and reconciliation by bilingual clinicians and researchers. It was pilot tested among 10 providers in the study region; pilot participants were excluded from the analytic cohort. Five knowledge indicators were analysed separately: three open-response disease-indication slots, Q18 (situations in which antibiotics are useful), and Q20 (an antibiotic not contraindicated in pregnancy). The two provisional attitude summaries averaged three equally weighted items each on a −2 to +2 response scale when at least two items were available: stewardship responsibility/self-efficacy (Q12, Q33, Q34) and AMR recognition/local salience (Q21, Q35, Q36). These summaries are treated as exploratory, provisional scores rather than validated scales. Analytic questionnaire items are listed in Supplementary Table S1; the complete bilingual questionnaire should accompany submission as a separate supplementary file.

### 2.6 Statistical Analysis

Descriptive statistics were first calculated for all provider variables of interest: counts and percentages for categorical variables and median, interquartile range (IQR), and range for age and years in practice. For hypothesis-directed provider analyses, the primary outcome was the four-level ordered cold/sore-throat antibiotic-frequency variable. Proportional-odds logistic regression was selected because the outcome categories have a natural order and the model estimates a single common odds ratio when the predictor effect is assumed constant across the cumulative thresholds. Thus, an adjusted common OR greater than 1 indicates greater odds of being in a higher prescribing-frequency category (for example, at least ‘some’ versus ‘none’, at least ‘most’ versus ‘some/none’, and ‘all’ versus lower categories) under the proportional-odds assumption. Models for guideline familiarity and medical-representative literature use adjusted for provider cadre, work setting, and years in practice. A binary any-versus-none logistic model was fitted as a sensitivity analysis to test whether conclusions were robust to collapsing the ordered outcome.

The proportional-odds assumption was assessed by likelihood-ratio comparison with an unconstrained multinomial-logit model and was supported for cold/sore-throat practice (p=0.592). In the broader separated-domain analysis, binary knowledge items were analysed with logistic regression, the two provisional attitude summaries with linear regression using HC3 robust standard errors, and ordered standalone items with proportional-odds logistic regression where the assumption was supported. Q22 and Q23 violated the assumption in their original five-level form and were therefore collapsed to disagree, neutral, and agree for sensitivity analyses. Outcome-specific complete-case analysis was used without imputation. Benjamini-Hochberg correction was applied within prespecified exploratory outcome families; because these multiplicity-adjusted domain analyses were secondary, the main text emphasizes effect estimates and confidence intervals rather than significance labels. All models converged; the maximum variance inflation factor was 2.83.

Provider analyses were performed in Python 3.12.13 using SciPy 1.17.0, NumPy 2.3.5, and pandas 2.2.3. The hypothesis-directed models were prespecified to address whether guideline familiarity or routine medical-representative literature use was associated with cold/sore-throat antibiotic frequency after adjustment for provider cadre, work setting, and experience. Throughout the manuscript, inferential effect estimates are reported together with their corresponding p-values; where Benjamini-Hochberg multiplicity correction was applied, the adjusted p-value (q) is also shown.

For the WHO/INRUD component, core indicators were summarized overall and by facility. For exploratory encounter-level correlates, antibiotic exposure (at least one antibiotic: yes/no) was the dependent variable. Each prescribing or care-process correlate was examined first without facility adjustment and then in a logistic model that included facility as a categorical adjustment factor. The manuscript reports group means/proportions together with effect estimates where available; these analyses evaluate association, not diagnosis-specific appropriateness. For exit interviews, categorical variables were summarized as counts and percentages and skewed expenditure by medians. Satisfaction (satisfied/very satisfied versus other responses) was modelled with multivariable binary logistic regression. Perceived clarity was modelled with proportional-odds logistic regression using provider type and education as predictors. Odds ratios are reported with 95% confidence intervals.

## 3. RESULTS

### 3.1 The Awareness-Practice Decoupling in Primary Care

#### 3.1.1 Provider Sample Characteristics

The 171 providers had a median age of 38 years (IQR 28-46; range 22-75). Of the full sample, 108 (63.2%) were male, 60 (35.1%) were female, and 3 (1.8%) had missing gender data. Provider cadres comprised 26 MMPs (15.2%), 65 AYUSH practitioners (38.0%), 33 allied health professionals (19.3%), and 47 informal practitioners (27.5%). Work setting was missing for 1 provider and years in practice for 1 provider. Descriptive characteristics are shown in Table 2a.

**Table 2a.** Provider sample characteristics (N=171).

| Characteristic | n / summary | % or distribution |
| --- | --- | --- |
| Age, years | 171 | Median 38; IQR 28-46; range 22-75 |
| Male | 108 | 63.2% |
| Female | 60 | 35.1% |
| Gender missing | 3 | 1.8% |
| MMP | 26 | 15.2% |
| AYUSH | 65 | 38.0% |
| Allied | 33 | 19.3% |
| Informal | 47 | 27.5% |
| Work setting missing | 1 | 0.6% |
| Years in practice missing | 1 | 0.6% |

AMR awareness was high across the provider cohort (91.4%, 149/163), and 95.8% (160/167) agreed that antibiotics are overused nationally. However, self-reported familiarity with guidelines was markedly lower, at 58.3% (95/163), dropping from 87.0% in MMPs to 48.9% in informal practitioners (p=0.014).

Despite high awareness, 80.4% (131/163) reported prescribing antibiotics for cold or sore throat in at least some patients. Routine use of medical-representative literature as a drug-information source was reported by 39.6% (65/164) overall: 53.2% among informal practitioners, 46.7% among AYUSH practitioners, 28.0% among MMPs, and 15.6% among allied providers. The between-cadre differences in this information-source variable were large in magnitude (Pearson chi-square p=0.0029); the p-value tests whether the distribution differs across the four cadres.

Table 2b reports outcome-specific denominators. Missing responses ranged from 4 (national-overuse belief) to 8 (AMR awareness and cold/sore-throat practice) overall and were retained as missing rather than included in cadre denominators.

**Table 2b.** Provider KAP indicators by cadre. Values are n/N (%) among non-missing responses.

| Cadre | Cadre sample (n) | AMR aware, n/N (%) | Knows guidelines, n/N (%) | Agrees overused, n/N (%) | Prescribes cold/ST, n/N (%) | Uses MR literature, n/N (%) |
| --- | --- | --- | --- | --- | --- | --- |
| MMP | 26 | 26/26<br>(100.0%) | 20/23<br>(87.0%) | 25/26<br>(96.2%) | 18/26<br>(69.2%) | 7/25<br>(28.0%) |
| AYUSH | 65 | 55/60<br>(91.7%) | 32/61<br>(52.5%) | 62/65<br>(95.4%) | 46/61<br>(75.4%) | 28/60<br>(46.7%) |
| Allied | 33 | 31/32<br>(96.9%) | 20/32<br>(62.5%) | 30/31<br>(96.8%) | 26/30<br>(86.7%) | 5/32<br>(15.6%) |
| Informal | 47 | 37/45<br>(82.2%) | 23/47<br>(48.9%) | 43/45<br>(95.6%) | 41/46<br>(89.1%) | 25/47<br>(53.2%) |
| Overall | 171 | 149/163<br>(91.4%) | 95/163<br>(58.3%) | 160/167<br>(95.8%) | 131/163<br>(80.4%) | 65/164<br>(39.6%) |
| Pearson chi-square p-value | - | 0.037 | 0.014 | 0.990 | 0.112 | 0.0029 |

#### 3.1.2 Adjusted Provider Analyses

Exploratory separated-domain models adjusted simultaneously for work setting, years in practice, provider cadre, and gender. For the stewardship responsibility/self-efficacy score, adjusted mean differences relative to MMPs were −0.32 (95% CI −0.59 to −0.06; BH-adjusted p [q]=0.036) for AYUSH, −0.53 (95% CI −0.82 to −0.25; BH-adjusted p [q]=0.001) for allied, and −0.55 (95% CI −0.79 to −0.31; BH-adjusted p [q]<0.001) for informal providers. These differences indicate lower mean provisional responsibility/self-efficacy scores in each of these cadres relative to MMPs; because the score is exploratory and not a validated scale, the magnitude should be interpreted cautiously.

For the provisional AMR recognition/local-salience score, adjusted mean differences relative to MMPs were −0.58 (95% CI −0.87 to −0.28; BH-adjusted p [q]=0.001) for allied providers and −0.46 (95% CI −0.70 to −0.22; BH-adjusted p [q]=0.001) for informal providers. Private work setting was also associated with lower mean responsibility/self-efficacy (−0.21, 95% CI −0.37 to −0.04; BH-adjusted p [q]=0.036) and lower recognition/local-salience (−0.24, 95% CI −0.44 to −0.04; BH-adjusted p [q]=0.036) than government work settings. These estimates are presented as exploratory effect sizes rather than as evidence of causal differences.

For cold/sore-throat practice, each additional 5 years in practice was associated with 1.36-fold higher odds of being in a higher antibiotic-provision frequency category (adjusted common OR 1.36, 95% CI 1.09-1.70; nominal p=0.006; BH-adjusted p [q]=0.041; n=158). Under the proportional-odds model, the same common OR applies across each cumulative threshold of the ordered outcome. The provider-cadre term also varied overall, but individual cadre contrasts were imprecise after accounting for multiple exploratory tests. Gender was not meaningfully associated with the ordered practice outcome.

**Table 2c.**
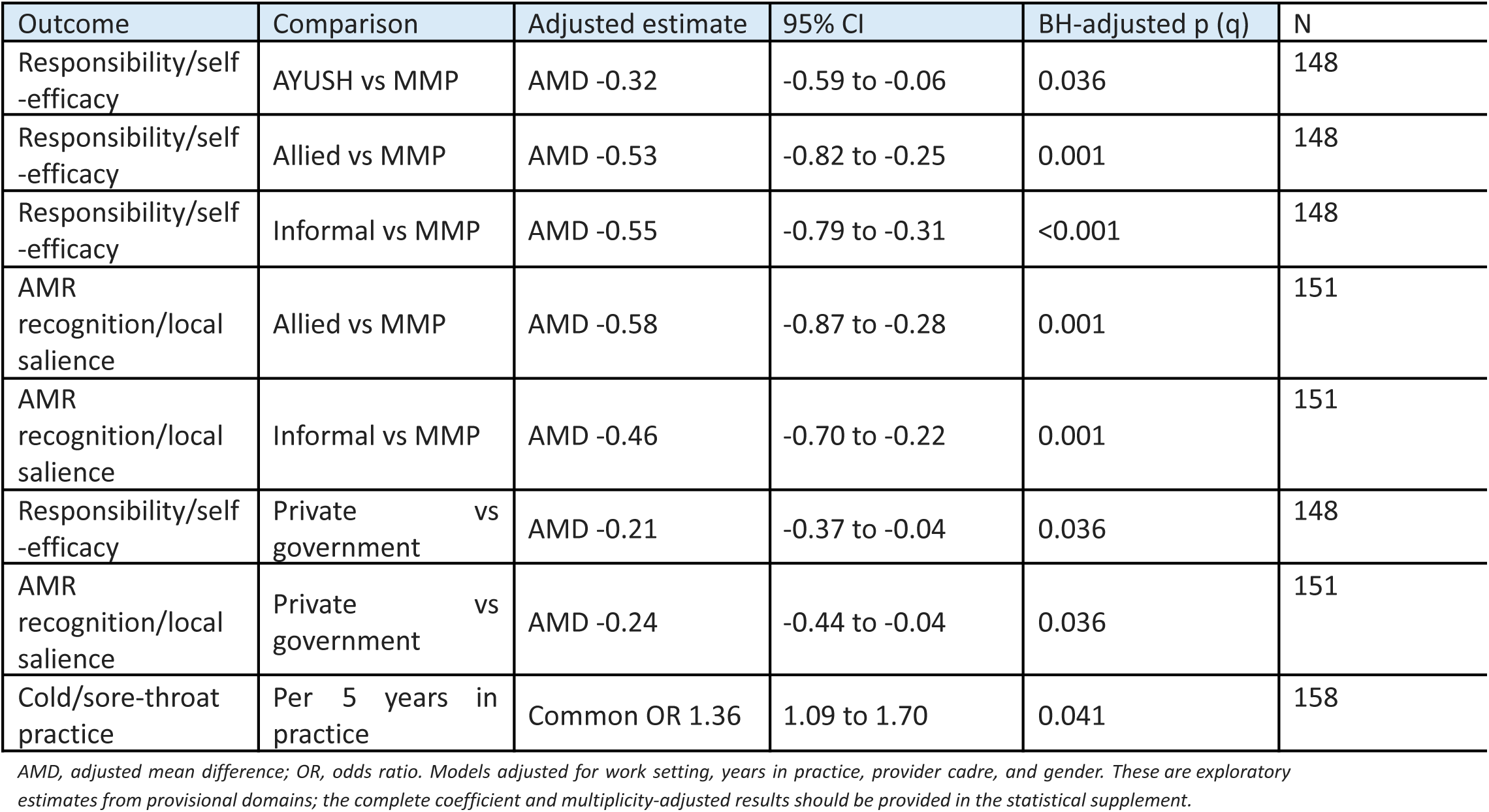
Selected adjusted estimates from exploratory separated-domain provider models.

In the primary hypothesis-directed proportional-odds model, guideline familiarity was not associated with lower cold/sore-throat antibiotic frequency (adjusted common OR 1.13, 95% CI 0.57-2.24; p=0.725). This common OR compares the odds of being in a higher versus lower prescribing-frequency category at each cumulative threshold, conditional on the proportional-odds assumption. The binary any-versus-none sensitivity model was concordant (adjusted OR 1.10, 95% CI 0.43-2.79; p=0.842).

Routine use of medical-representative literature was associated with higher estimated prescribing frequency, but the confidence interval included both little association and a potentially important increase (adjusted common OR 1.89, 95% CI 0.88-4.07; p=0.104). Informal-practitioner coefficients in the two hypothesis-directed models were 4.17 (95% CI 1.29-13.50; p=0.017) and 3.79 (1.21-11.94; p=0.023) relative to MMPs. These informal-provider contrasts were exploratory and should be interpreted alongside the broader cadre analysis rather than as confirmatory tests.

**Table 2d.**
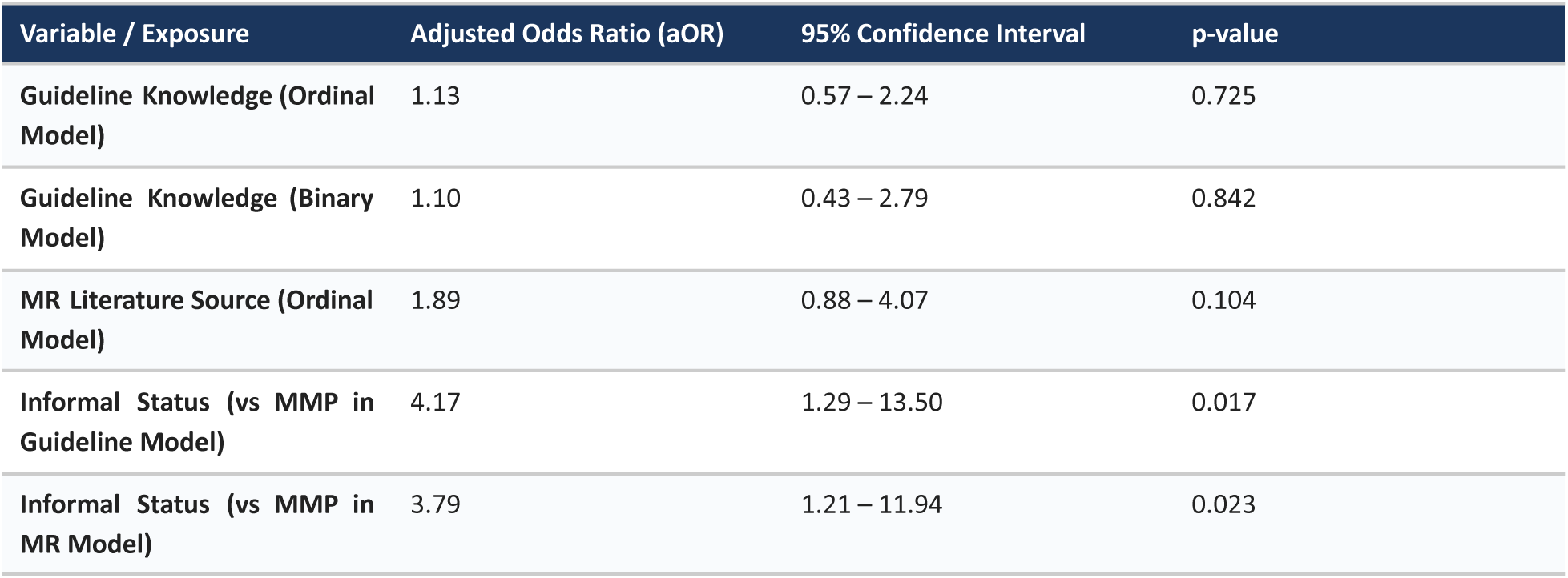
Hypothesis-directed models for cold/sore-throat antibiotic frequency.

| Variable / Exposure | Adjusted Odds Ratio (aOR) | 95% Confidence Interval | p-value |
| --- | --- | --- | --- |
| Guideline Knowledge (Ordinal Model) | 1.13 | 0.57 – 2.24 | 0.725 |
| Guideline Knowledge (Binary Model) | 1.10 | 0.43 – 2.79 | 0.842 |
| MR Literature Source (Ordinal Model) | 1.89 | 0.88 – 4.07 | 0.104 |
| Informal Status (vs MMP in Guideline Model) | 4.17 | 1.29 – 13.50 | 0.017 |
| Informal Status (vs MMP in MR Model) | 3.79 | 1.21 – 11.94 | 0.023 |

### 3.2 Facility Prescribing Norms and Polypharmacy

Across 600 audited encounters in 20 government primary healthcare facilities, 54.8% (329/600) included at least one antibiotic, and 11.8% (71/600) contained an injection. Overall generic prescribing was 72.8%, and EDL concordance was 50.0%. Facility-level variation was substantial: antibiotic prescribing ranged from 16.7% to 80.0% across facilities, generic prescribing ranged from 32.6% to 100%, and EDL compliance ranged from 32.6% to 94.7%. These indicators quantify prescribing exposure and composition, not diagnosis-matched appropriateness.

At the encounter level, antibiotic-containing prescriptions had more medicines (mean 2.88 vs 2.47), lower generic proportions (73.6% vs 80.7%), lower EDL concordance (48.9% vs 63.8%), and more co-prescribed injections (15.2% vs 7.7%). In unadjusted logistic models with antibiotic exposure as the dependent variable, these differences corresponded to p<0.001, p=0.003, p<0.001, and p=0.007, respectively. These p-values test association with antibiotic exposure; the group estimates convey the magnitude of the differences.

After adding facility as a categorical adjustment factor, total drug count remained associated with antibiotic exposure (OR 1.52 per additional medicine; p<0.001), and the inverse association with generic prescribing remained (p=0.002). The EDL association attenuated (p=0.066), as did co-prescribed injection (OR 2.33; p=0.058). Consultation-time and dispensing-time associations also attenuated after facility adjustment (p=0.051 and p=0.541, respectively). Because diagnoses were unavailable, none of these analyses establishes whether an antibiotic prescription was clinically appropriate.

Patient-care indicators showed very short clinical encounters despite generally high medicine-dispensing performance. Mean consultation time was 106.8 seconds (1.78 minutes) and mean dispensing time was 62.0 seconds. Overall, 93.4% of prescribed medicines were dispensed and 89.1% of dispensed medicines were adequately labelled. Recorded patient knowledge of the correct dosage was 99.8%. Considerable between-facility variation was present, particularly for consultation and dispensing times and adequate labelling.

**Table 3.**
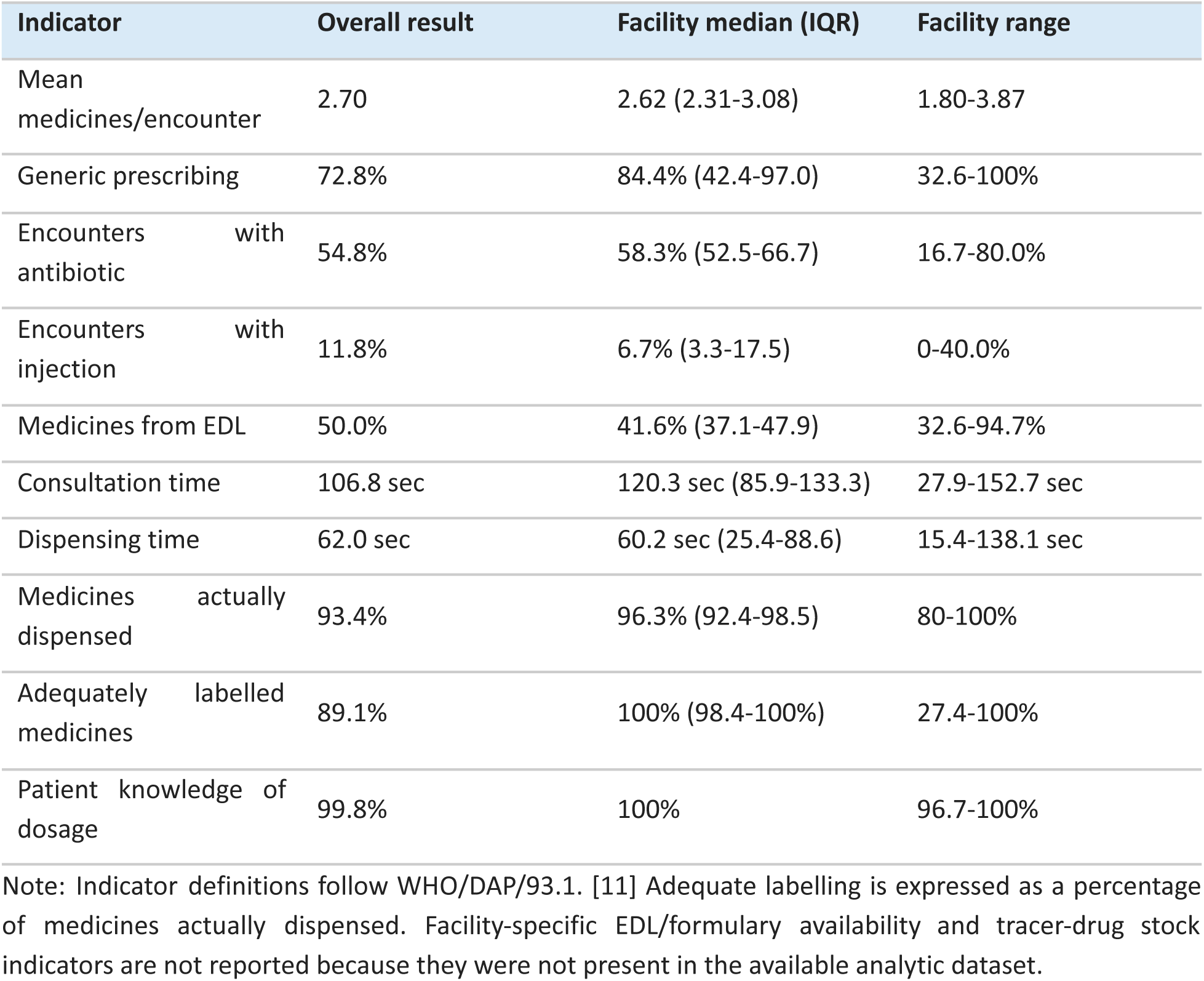
WHO/INRUD prescribing and patient-care indicators with facility variation.

| Indicator | Overall result | Facility median (IQR) | Facility range |
| --- | --- | --- | --- |
| Mean medicines/encounter | 2.70 | 2.62 (2.31-3.08) | 1.80-3.87 |
| Generic prescribing | 72.8% | 84.4% (42.4-97.0) | 32.6-100% |
| Encounters with antibiotic | 54.8% | 58.3% (52.5-66.7) | 16.7-80.0% |
| Encounters with injection | 11.8% | 6.7% (3.3-17.5) | 0-40.0% |
| Medicines from EDL | 50.0% | 41.6% (37.1-47.9) | 32.6-94.7% |
| Consultation time | 106.8 sec | 120.3 sec (85.9-133.3) | 27.9-152.7 sec |
| Dispensing time | 62.0 sec | 60.2 sec (25.4-88.6) | 15.4-138.1 sec |
| Medicines actually dispensed | 93.4% | 96.3% (92.4-98.5) | 80-100% |
| Adequately labelled medicines | 89.1% | 100% (98.4-100%) | 27.4-100% |
| Patient knowledge of dosage | 99.8% | 100% | 96.7-100% |

Note: Indicator definitions follow WHO/DAP/93.1. [11] Adequate labelling is expressed as a percentage of medicines actually dispensed. Facility-specific EDL/formulary availability and tracer-drug stock indicators are not reported because they were not present in the available analytic dataset.

### 3.3 Sector-Specific Market Dynamics and Parenteral Exposure

In the broader market sample of 371 patient exit interviews, 48.8% (181/371) reported receiving an injection. Parenteral exposure was concentrated within the informal sector: 92.3% (131/142) of informal provider encounters involved an injection. Consequently, informal practitioners accounted for ∼72.4% (131/181) of all reported injections in the marketplace despite representing 38.3% of consultations. Injection administration was reported in 7.5% of government encounters, closely mirroring the 11.8% rate observed in the independent government WHO audit. The data did not identify the contents of the injections or establish whether the injections were clinically unnecessary or administered unsafely. Median out-of-pocket expenditure was INR 0 at government facilities, INR 18 at NGO facilities, INR 150 at informal providers, and INR 200 at private settings.

**Table 4.**
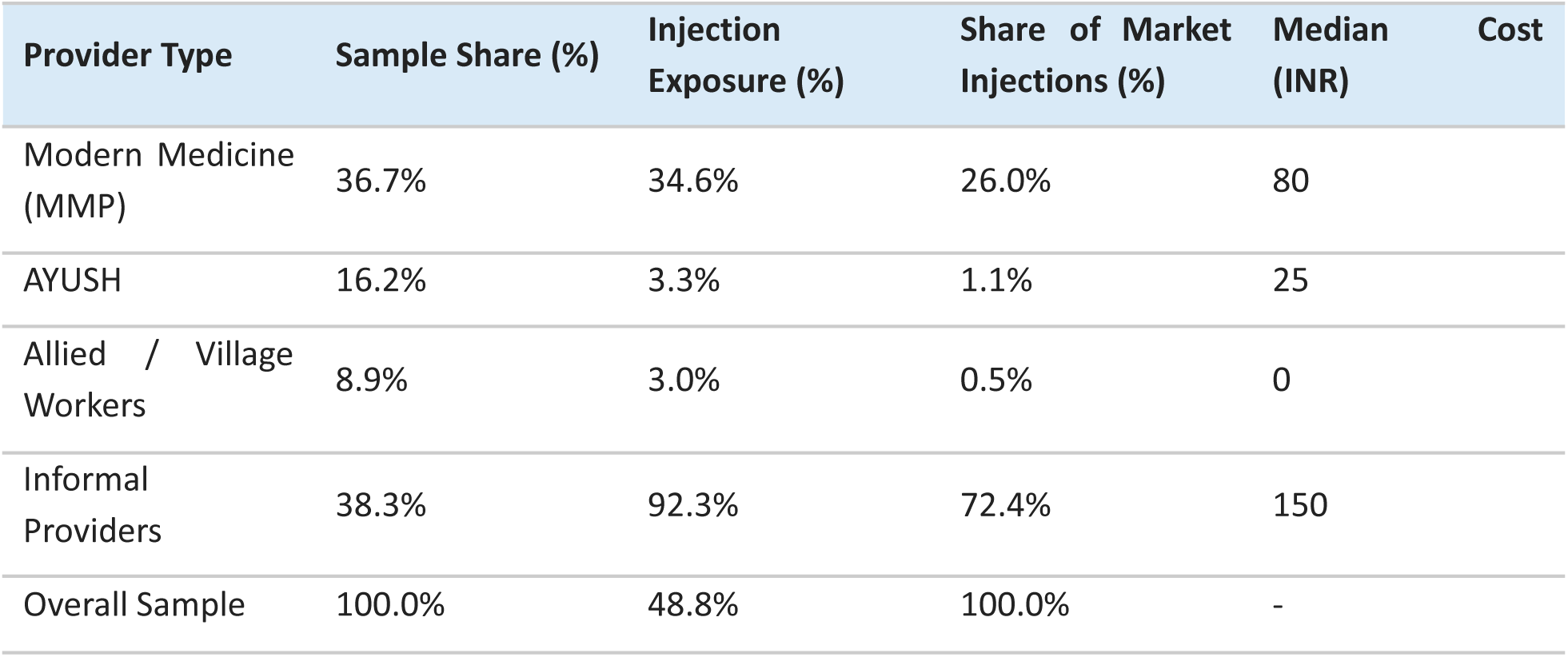
Exit-Interview Market Indicators by Provider Type.

### 3.4 Patient Experience and Determinants of Satisfaction

Overall, 281/371 (75.7%) respondents were satisfied or very satisfied. In the complete-case multivariable binary logistic model (n=369), clear or very clear instructions were strongly associated with satisfaction (adjusted OR 13.66, HC3 95% CI 6.73-27.71; p<0.001). The model also included written-prescription receipt, injection exposure, out-of-pocket expenditure, education, and facility sector. Because satisfaction and clarity were measured in the same interview, the large association may partly reflect construct overlap or common-method variance.

Perceived clarity was modelled on its five ordered response levels. Relative to MMP encounters, the adjusted odds of a higher clarity rating were lower for informal providers (adjusted common OR 0.28, 95% CI 0.17-0.46; p<0.001) and village health workers (0.48, 0.23-0.98; p=0.045), and higher for AYUSH practitioners (2.05, 1.16-3.63; p=0.013). Patient education was not associated with clarity (adjusted common OR 0.94 per education category, 95% CI 0.79-1.13; p=0.531).

## 4. STEWARDSHIP IMPLICATIONS AND EVIDENCE-TO-ACTION MATRIX

By triangulating provider KAP, government-facility prescribing assessments, and patient exit interviews, this study uses a diagnostic approach to identify which stewardship barriers are most salient in the three study districts. The intended transferable contribution is this approach to context assessment, not the assumption that the same prevalence estimates or dominant mechanisms apply elsewhere. The proposed responses align with India’s National Action Plan on AMR 2.0 and WHO AWaRe guidance, but implementation should be adapted to local provider mix, facility context, information environment, and patient-care market. Injection findings also require a distinct infection-prevention and injection-safety pathway. [12–15]

**Table 5.**
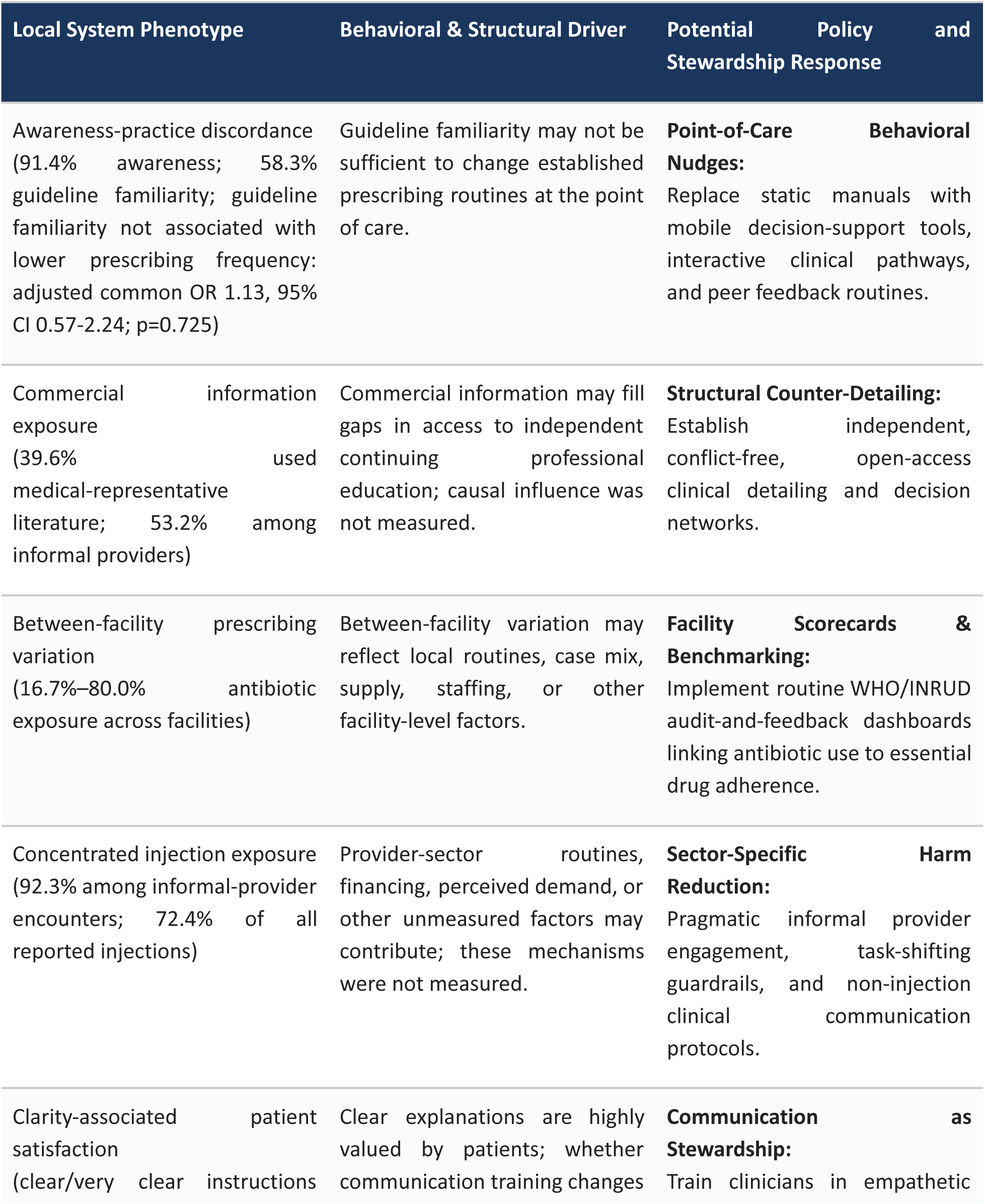
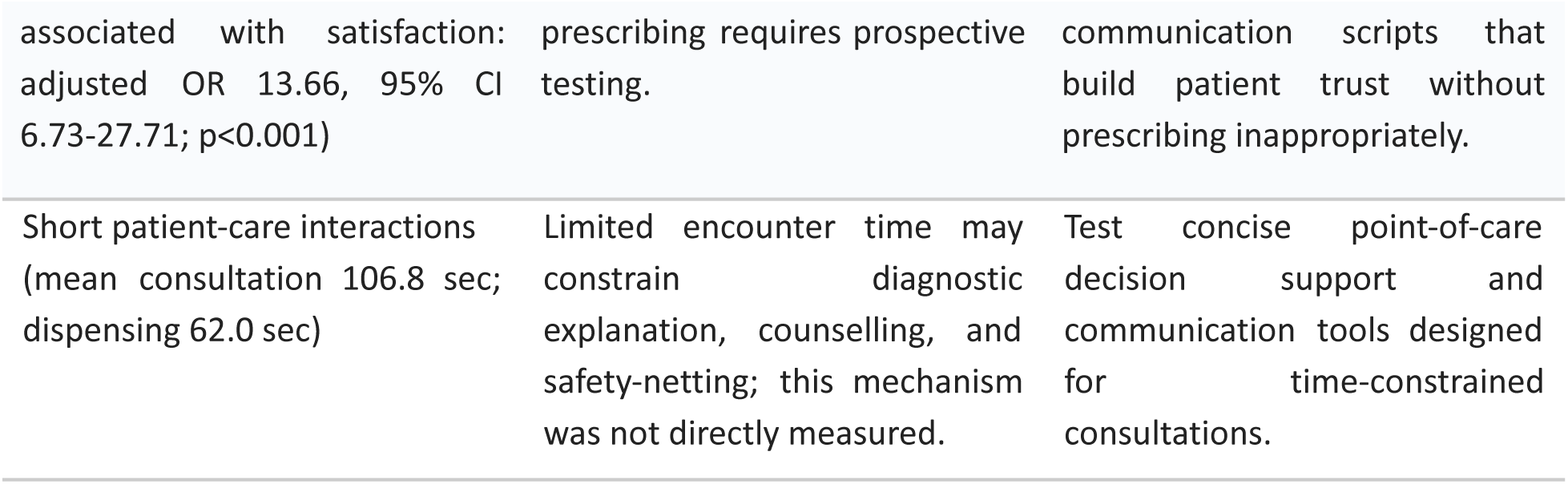
Evidence-to-action matrix for stewardship interventions requiring prospective evaluation.

### 4.1 Behavioral Interpretation and Testable Intervention Targets

The three components were not linked at the individual level. Accordingly, the framework below separates observations directly supported by the data from hypothesized mechanisms and interventions that require prospective evaluation. The interpretation is organized around the capability, opportunity, and motivation domains commonly used in behavior-change research.

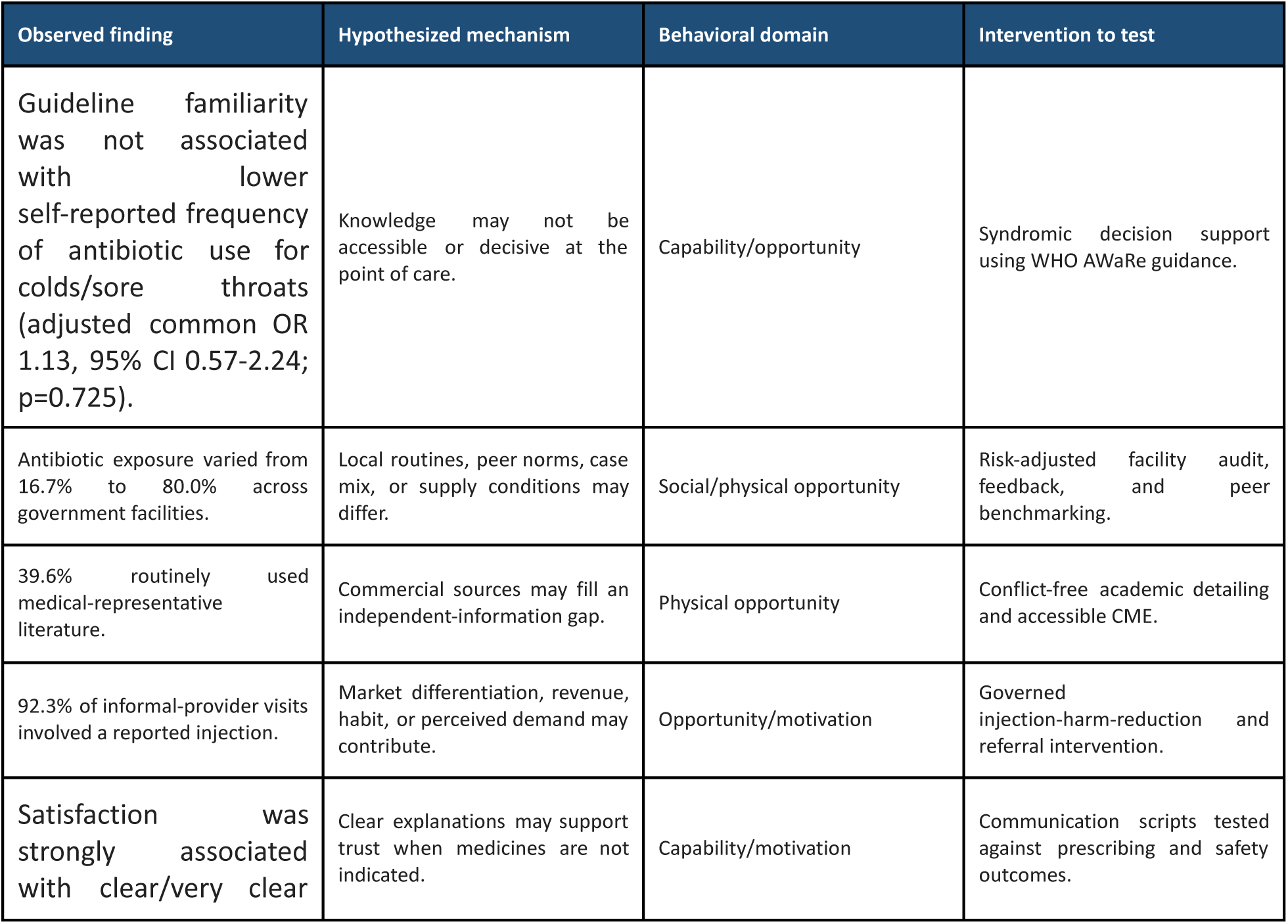
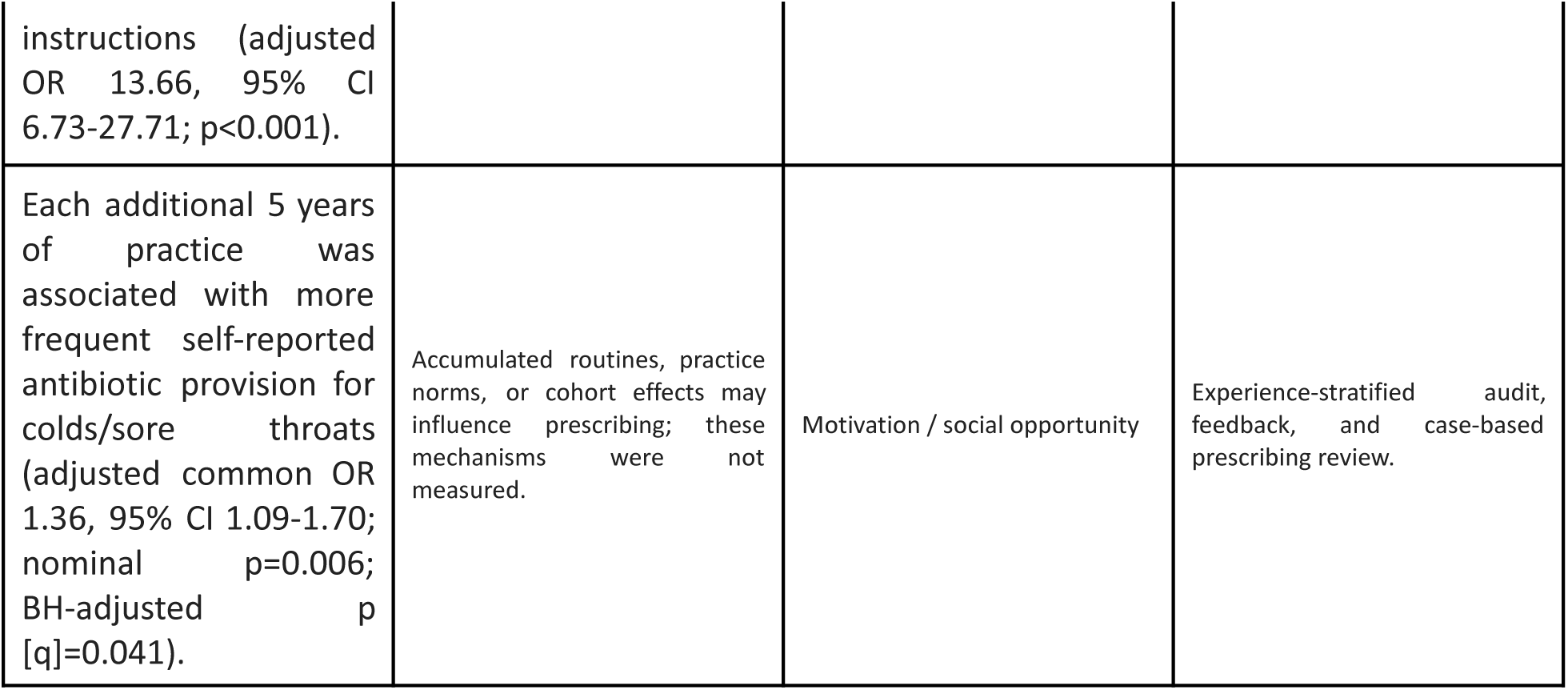

## 5. DISCUSSION

### 5.1 Beyond Knowledge Dissemination in Antimicrobial Stewardship

AMR programmes often include education and guideline dissemination. In this study, 95.8% of responding providers agreed that antibiotics are overused in India, yet 80.4% reported providing antibiotics for cold or sore throat to at least some patients. Guideline familiarity was not associated with lower self-reported prescribing frequency (adjusted common OR 1.13, 95% CI 0.57-2.24; p=0.725). The estimate is compatible with no meaningful protective association in this sample.

Exploratory provider-domain analyses suggest that provider background and accumulated practice experience may be relevant to intervention design. Each additional 5 years in practice was associated with higher odds of being in a more frequent cold/sore-throat antibiotic-provision category (adjusted common OR 1.36, 95% CI 1.09-1.70; nominal p=0.006; BH-adjusted p [q]=0.041). Differences in the provisional attitude scores were also observed across cadres, but these scores had only moderate internal consistency and were not validated for cross-cadre comparisons. They should therefore be viewed as descriptive signals rather than definitive latent constructs.

These findings are consistent with, but do not demonstrate, a role for prescribing routines, diagnostic uncertainty, peer norms, and workplace context in addition to knowledge. The principal transferable contribution of this study is therefore not the observed prevalence of any single prescribing behavior in Chhattisgarh. Rather, it is the diagnostic approach: before stewardship interventions are implemented, locally generated evidence can be used to identify which behavioral, organizational, informational, and sectoral determinants are most relevant in a given health system. In the three districts studied, a reasonable next step is to prospectively test whether point-of-care decision support, audit and feedback, peer comparison, independent drug information, or targeted informal-provider engagement changes prescribing. Applicability elsewhere in Chhattisgarh, India, or other countries should be evaluated rather than assumed.

### 5.2 Patient-Care Indicators and Stewardship Opportunity

The WHO/INRUD patient-care indicators identify additional system-level stewardship constraints. Consultations averaged less than two minutes and dispensing interactions approximately one minute, potentially limiting opportunities for diagnostic assessment, explanation of non-antibiotic management, counselling, and safety-netting. At the same time, most prescribed medicines were dispensed and recorded dosage knowledge was nearly universal. The latter finding should be interpreted cautiously because its near-constant distribution may reflect the way the indicator was recorded rather than uniformly comprehensive counselling. These patient-care indicators were defined according to WHO/DAP/93.1. [11]

### 5.3 Countering Commercial Dominance in Decentralized Markets

In the study sample, 39.6% of providers used medical-representative literature as a routine source of drug information, including 53.2% of informal practitioners. Continuing medical education (CME) and other independent information sources may be less accessible in some decentralized settings, but this study did not measure why providers used commercial literature. The adjusted association between medical-representative literature use and cold/sore-throat antibiotic frequency was directionally positive but imprecise (adjusted common OR 1.89, 95% CI 0.88-4.07; p=0.104). This supports testing independent academic-detailing approaches, not a causal claim about pharmaceutical marketing.

### 5.4 Pragmatic Governance of the Informal Sector

In the three study districts, informal providers represented an important part of the observed care market. The exit-interview findings support targeted safety-oriented engagement because informal providers accounted for 72.4% of all reported injections and 92.3% of their encounters involved an injection. The study did not measure whether those injections were clinically indicated or unsafe, so proposed injection-reduction or referral interventions should be evaluated prospectively and within existing regulatory and scope-of-practice frameworks.

Because unnecessary or unsafe parenteral administration can create infection-prevention, blood-borne pathogen, and health-care waste risks, stewardship programs must prioritize informal providers through carefully governed, scope-appropriate interventions: injection-reduction campaigns, basic diagnostic safety, and communication scripts for symptomatic non-pharmacological care.

### 5.5 Communication as an Antimicrobial Stewardship Intervention

Clinicians may perceive that patients value active treatment, but this exit-interview study did not directly measure antibiotic demand. It did show that reported receipt of an injection was not the dominant correlate of satisfaction, whereas clear or very clear instructions were strongly associated with satisfaction (adjusted OR 13.66, 95% CI 6.73-27.71; p<0.001).

This association identifies communication as a candidate stewardship lever. Structured explanations of expected illness course, symptomatic care, antibiotic non-indication, and return precautions could be tested for effects on prescribing, revisits, safety, and patient trust. The present cross-sectional data do not establish that communication training will reduce antibiotic or injection use.

Our findings suggest that stewardship in heterogeneous health systems requires ground-level assessment before implementation. The transferable lesson is the principle of context adaptation rather than the exact frequencies observed in this study. Prescribing behavior varied by provider cadre and facility, commercial information sources were common, and injection exposure was concentrated in the informal sector. These patterns indicate that interventions imported from more uniformly regulated health systems may not address the dominant local behavioral and structural drivers. WHO and national guidance should therefore serve as an evidence framework, while the mode of implementation is adapted to the local care market, tested prospectively, and monitored for effectiveness and unintended effects.

### 5.6 Strengths and Limitations

A key strength is the use of three complementary perspectives: provider-reported knowledge and practice, government-facility prescribing indicators, and patient-reported market experience. Their independence reduces the likelihood that a single measurement method can explain every pattern, but it also prevents person- or facility-level linkage. The observational, cross-sectional design precludes causal attribution. Provider responses may be affected by recall and social-desirability bias; prescription audits did not include diagnoses, indications, antibiotic class, dose, or duration; and exit interviews did not identify injection contents or safety practices. Differential case mix, severity, access, supply conditions, and unmeasured provider or facility characteristics may explain part of the observed differences. Calendar period was partly confounded with facility identity. The large clarity–satisfaction association may also reflect common method variance or construct overlap. Finally, the study was conducted in three districts of a single Indian state; transferability to other settings depends on workforce regulation, financing, pharmaceutical markets, and service availability. The findings should therefore be interpreted as system-level diagnostic signals and intervention hypotheses rather than estimates of causal effects. The separated KAP constructs had only moderate internal consistency and were not demonstrated to be measurement-invariant across cadres; they should therefore be interpreted as provisional construct scores rather than validated scales. Complete-case sample sizes ranged from 100 to 166, largely due to progressive missingness in the open-response disease slots. Q22 and Q23 violated the five-level proportional-odds assumption, and only collapsed sensitivity models should inform their adjusted interpretation. The cold/sore-throat measure was a single self-reported practice indicator and did not establish diagnosis-matched prescribing inappropriateness.

## 6. POLICY RECOMMENDATIONS FOR THE STUDY SETTING AND PROSPECTIVE EVALUATION

1. Pivot from Passive Guidelines to Active Decision Nudges: Complement passive guideline dissemination with locally adapted point-of-care pathways based on WHO AWaRe guidance, and evaluate uptake, antibiotic selection, safety, and equity before scale-up.
2. Establish Institutional Audit & Peer-Benchmarking Dashboards: Pilot monthly facility scorecards that report antibiotic exposure, polypharmacy, essential-medicine concordance, and—where drug names are available—AWaRe composition; use risk-aware peer comparison and monitor unintended effects.
3. Deploy Targeted Parenteral Harm Reduction: Co-design governed injection-harm-reduction initiatives with informal-provider networks, emphasizing injection minimization, single-use safety, referral, and scope-of-practice safeguards without conferring unrestricted prescribing authority.
4. Institutionalize Prescriber-Patient Communication Training: Test structured communication training that explains expected illness course, symptomatic care, antibiotic non-indication, and return precautions while measuring prescribing, revisits, adverse outcomes, and patient trust.

## SUPPLEMENTARY MATERIAL

**Supplementary Table S1.** Provider questionnaire items used in the analyses reported in this manuscript.

| Variable name | Question / construct | Response used | Role in analysis |
| --- | --- | --- | --- |
| AMR awareness | Awareness/recognition of antimicrobial resistance | Yes/no or prespecified response | Descriptive |
| National overuse belief | Q21: Antibiotics are overused in India | Agreement scale | Descriptive; component of provisional recognition score |
| Guideline familiarity | Knowledge of any guideline related to antibiotic use | Yes/no | Primary hypothesis-directed exposure |
| MR literature source | Usual drug-information source includes literature from medical representatives | Yes/no | Primary hypothesis-directed exposure |
| Cold/sore-throat practice | Q27: Frequency of prescribing/providing an antibiotic for a cold or sore throat | None / Some / Most / All | Primary ordered outcome |
| Disease-indication knowledge 1-3 | Top three diseases for which antibiotics are required | Open response; prespecified scoring | Exploratory knowledge outcomes |
| Q18 knowledge | Situations in which antibiotics are useful | Prespecified scoring | Exploratory knowledge outcome |
| Q20 pregnancy safety | Antibiotic not contraindicated in pregnancy | Prespecified scoring | Exploratory knowledge outcome |
| Responsibility/self-efficacy | Mean of Q12, Q33, Q34 when answered $\geq 2$ | -2 to +2 | Exploratory provisional score |
| AMR recognition/local salience | Mean of Q21, Q35, Q36 when answered $\geq 2$ | -2 to +2 | Exploratory provisional score |
| Patient-demand pressure | Q22: Patient demands contribute to antibiotic overuse | Agreement scale | Exploratory standalone item |
| Harm recognition | Q23: Prescribing when not indicated is harmless (reverse-scored) | Agreement scale | Exploratory standalone item |
| Education receptivity | Q24: Would like access to educational programmes on antibiotic prescribing | Agreement scale | Exploratory standalone item |

Note: This table documents the analytic items used in the manuscript. The complete original Hindi-English questionnaire should be submitted as a separate supplementary file because the reviewed manuscript did not contain every instrument item.

## DECLARATIONS

**Ethics Approval and Consent to Participate:** This study was approved by the Shaheed Hospital Institutional Ethics Committee (SHD/26/2020; May 9, 2020). Informed consent was obtained from all participants before enrolment.

**Funding:** Funded by the Indian Council of Medical Research (ICMR; Grant AMR/194/2019/EDC-II). The funder had no role in study design, data collection, analysis, interpretation, or writing of the report.

**Competing Interests:** The authors declare no competing interests.

**Data Availability:** Cleaned analytic datasets and data dictionaries are available from the corresponding author upon reasonable request.

## Data Availability

Cleaned analytic data and data dictionaries are available from the corresponding author upon reasonable request.

